# Anti-oxidative stress therapies reduce chemotherapy-induced peripheral neuropathy in colorectal cancer patients treated with oxaliplatin: a systematic review and meta-analysis

**DOI:** 10.1101/2025.03.24.25324521

**Authors:** Mohamed Salama, David Barnes, Anni Georghiou, Mariam Murad, Seham Almalki, Zubair Ahmed, Mark R Openshaw, Claire Palles, Richard I Tuxworth

**Affiliations:** Birmingham Centre for Neurogenetics, University of Birmingham, Birmingham, B15 2TT; Department of Cancer and Genomic Sciences, School of Medical Sciences, University of Birmingham, Birmingham B15 2TT; Department of Inflammation and Ageing, School of Infection, Immunity and Inflammation, University of Birmingham, Birmingham, B15 2TT; Birmingham NIHR Biomedical Research Centre, University of Birmingham, Birmingham, B15 2TT; Department of Biotechnology, Faculty of Science, Taif University, Taif, Saudi Arabia; University Hospitals Birmingham NHS Foundation Trust, Mindelsohn Way, Edgbaston, Birmingham, B15 2GW

**Keywords:** bowel cancer, CIPN, platinum agents, neurotoxicity, oxidative stress, mitochondrial dysfunction

## Abstract

**Background and Purpose:** Chemotherapy-induced peripheral neuropathy (CIPN) is a major side-effect of many commonly used cancer drugs, affecting up to 90% of patients treated with oxaliplatin. This systematic review and meta-analysis aimed to analyse randomised controlled trials (RCTs) to determine if any pharmacological agents can prevent or reduce oxaliplatin-induced peripheral neuropathy (OIPN) in colorectal cancer (CRC) patients.

**Materials and Methods:** We searched PubMed, EMBASE and Web of Science for RCTs published before March 2025 that included patients with CRC who received oxaliplatin-based chemotherapy and had peripheral neuropathy quantified using Common Toxicity Criteria for Adverse Events (CTCAE). Meta-analysis was performed for agents tested in three or more RCTs with a minimum combined sample size of 100 patients.

**Results:** 20 studies were included with a median sample size of 61 (range 14-2450). Meta-analysis was used for two treatments. Anti-oxidative stress treatments were not associated with reduced incidence of grade ≥2 OIPN when assessed mid-treatment but were associated with a significant reduction of grade ≥2 OIPN at the end of treatment (OR:0.04, 95%CI:0.01-0.12; p<0.00001). No reduction of grade ≥2 OIPN was observed for Ca^2+^/Mg^2+^ infusions. 35% of studies had potential high risk of bias and 45% of studies showed low risk of bias.

**Conclusions:** Whilst the existing published RCTs included small numbers of patients, the meta-analysis indicates that anti-oxidative stress therapies reduce the end of treatment severe OIPN incidence in CRC patients. A large, randomised, placebo-controlled trial assessing OIPN using CTCAE grades and patient-reported outcomes is warranted to confirm these findings.

## Introduction

Chemotherapy-induced peripheral neuropathy (CIPN) is one of the most frequent side-effects of commonly used anti-neoplastic agents, including platinum drugs, taxanes and proteosome inhibitors. 30-50% of patients experience CIPN with low dose regimens but at higher doses as many as 90% of patients experience neuropathy making it an important dose limiting toxicity.^1^ Symptoms include pain in the fingers and toes, loss of sensation, cold or mechanical allodynia and mechanical weakness. Symptoms can be life long, and in severe cases life changing. Motor symptoms are common, and many patients also report long-lasting cognitive effects. Neurotoxicity is severe in many cases, forcing tapering of chemotherapy regimens or even cessation of treatment, thereby limiting the efficacy of cancer treatment.^2^

Oxaliplatin, a third-generation platinum compound, is one of the most commonly used chemotherapeutic agents for treating colorectal cancer (CRC).^3^ It can cause acute neuropathy during, or immediately after infusion and symptoms of oxaliplatin-induced peripheral neuropathy (OIPN) can also emerge weeks or months later after the completion of chemotherapy.^4^ The severity of symptoms is usually proportional to the cumulative dose of the drug and therefore progressively worsens during therapy.^5^ A recent meta-analysis in patients treated for CRC found that 58% of patients reported OIPN 6 months after treatment, 45% at 12 months, 32% at 24 months, and 24% at 36 months.^6^ Patients treated with oxaliplatin often exhibit a coasting phenomenon in which OIPN symptoms continue to worsen for several months after treatment.^7^ Platinum drugs generate DNA damage in the nucleus but mitochondria are also affected and several studies have suggested mitochondrial damage is central to OIPN.^8^

CRC is now the third most common cancer in the UK and the second most common in males aged 45 to 74 years and females aged 45 to 54 years.^9^ Oxaliplatin-based treatment is the standard adjuvant treatment for stage III CRC and is a standard first line treatment for metastatic CRC patients. Hence, there is an urgent need to find therapies to prevent or reduce the severity of OIPN. No current therapies are effective at preventing or reducing the severity of OIPN; duloxetine, venlafaxine, gabapentin, pregabalin, lamotrigine, and amitriptyline are commonly given as initial treatments for neuropathic pain, but these drugs are only of limited benefit.^10^

Three systematic reviews have previously been published examining the evidence for the utility of pharmacological interventions in reducing incidence of peripheral neuropathy induced by chemotherapy agents. Two included studies treating cancer patients with any chemotherapeutic agent^11,12^ and one focused specifically on studies treating with oxaliplatin.^13^ Hershman *et al*, 2014^11^ reviewed studies published before April 2013, and Loprinzi *et al*, 2020^12^ reviewed studies published between 2013 and 2020. Both concluded that there are no pharmacological interventions that can currently be recommended to prevent CIPN. Hershman *et al.* concluded that there was strong evidence to recommend against the use of acetyl-L-carnitine, diethyldithiocarbamate, nimodipine and moderate evidence against using amifostine, amitriptyline, Ca^2+^ and Mg^2+^ infusions, Org 2766, all-trans retinoic acid, recombinant human leukaemia inhibitory factor (rhuLIF) and vitamin E, or glutathione in paclitaxel/carboplatin-treated patients only. Loprinzi *et al*. found a lack of evidence of benefit for use of calmangafodipir, cannabinoids, carbamazepine, L-carnosine, gabapentin/pregabalin, glutamate, goshajinkigan, metformin, minocycline, N-acetylcysteine, omega-3 fatty acids, oxcarbazepine, recombinant human leukaemia inhibitory factor, venlafaxine, vitamin B and E.

Peng *et al*, 2022^13^ restricted their review to trials of oxaliplatin therapy published before August 2020; trials of multiple cancer types were included. The evidence for 29 pharmacological interventions aiming to reduce OIPN was reviewed and 2 interventions were subsequently analysed by meta-analyses of 2 and 3 studies, respectively: N-acetylcysteine and glutathione. Both treatments were associated with a lower risk of common toxicity criteria (CTCAE) grade ≥2 OIPN. However, of the 5 studies included in the meta-analyses, 4 were assessed as having unclear or high risk of bias, making interpretation difficult. Peng *et al*.^13^ also noted substantial differences in timing and/or scoring methods used to assess the severity of OIPN between trials, and that many trials were not double-blind, randomised, placebo-controlled, all of which make analysis difficult or impossible.

Here, we report the results of a new systematic review of pharmacological interventions assessed for their ability to reduce OIPN in a clinical trial including CRC patients and published before March 2025. Among other strict inclusion criteria, we only analysed studies that had assessed OIPN using the CTCAE scale and we performed sensitivity analyses excluding trials where it was unclear if blinding had been applied. Meta-analysis was only performed for interventions investigated by three or more independent studies.

## Materials and methods

### Search strategy

We searched PubMed, EMBASE and Web of Science in March 2025 for peer-reviewed English-language randomised controlled trials (RCTs). Searches were not limited by date restrictions. Search terms were: “Oxaliplatin induced peripheral neuropathy treatment” OR “Oxaliplatin induced neurotoxicity treatment” OR “Chemotherapy induced peripheral neuropathy and therapy” OR “Chemotherapy induced neurotoxicity and therapy”. Additional references were also obtained from the references cited by a recent relevant meta-analysis.^13^ Two investigators (M.S. and Z.A.) independently read and selected the retrieved abstracts. Discrepancies between the reviewers’ selections were resolved by discussion. Full text versions of potentially eligible studies were then read by M.S. to confirm each conformed to the inclusion and exclusion criteria.

### Inclusion criteria

RCTs were included if worst OIPN grade on treatment in adult CRC patients was evaluated using the CTCAE scale and patients were randomised to a pharmacological agent or traditional herbal medicine being tested as a preventative treatment for OPIN. To be included in the meta-analysis, studies needed to report how many participants in each arm had experienced either <grade 2 OIPN or ≥grade 2 OIPN and specify the time point at which this was measured.

### Exclusion criteria

Studies were excluded if: there was no full text published in English; OIPN was only evaluated using an alternative to the CTCAE scale; it was not possible to calculate counts <grade 2 and ≥grade 2 OIPN (n=4);^14,3,15,16^ analysis included patients with baseline peripheral neuropathy (PN) (n=2);^17,18^ the study aimed to treat rather than prevent PN (n=2);^19^ patients with ECOG performance status 3 were included in the analysis (n=1);^20^ or ≤10 patients were available for analysis (n=1).^21^

### Timepoints when OIPN was assessed

Different trials assessed OIPN at different time points. Where available, we extracted data from trials that assessed OIPN grade midway through treatment (cycles 4-6) and at the end of treatment (cycles 8-12) separately. In the Ca^2+^/Mg^2+^ infusion trials,^22,23,24,25^ highest PN CTCAE grades on treatment were provided. We combined results for CTCAE grade 2 and above since grade 2 is the usual threshold for clinical interventions, such as dose reduction of chemotherapy or treatment deferral.

### Data extraction and quality assessment

Data was extracted from the 20 included studies by two of 5 investigators independently (M.S., M.M, D.B., A.G. and C.P.). Data was imported into RevMan.^26^

### Meta-analysis

Inverse weighted random effects meta-analysis was performed in RevMan^26^ for pharmacological agents or classes of drugs if there were data from a minimum of 100 patients across 3 or more studies. Heterogeneity was estimated using the restricted maximum likelihood method. Risk of bias was also performed using RevMan, assisted by the RoB-2 tool.^27^

## Results

### Study selection and included interventions

Figure 1 shows the PRISMA flow chart of how RCTs were identified and reviewed. We reviewed the full-text report of 29 studies (Supplementary Table 1), of which 20 met the inclusion criteria (summarised in Table 1 with full details supplied in Supplementary Table 2). 12 studies were double-blinded and placebo controlled;^28,29,22,23,24,25,30,31,32,33,34,35^ four were placebo controlled but blinding was unclear;^36,37,38,39^ and four studies compared to a control arm with no or unclear blinding.^40,41,42,43^ Median incidence of grade ≥2 OIPN, as measured at the end of treatment (8-12 cycles), was 57.95% (range 31.2-100%) in the placebo or control arms.

**Figure 1.**
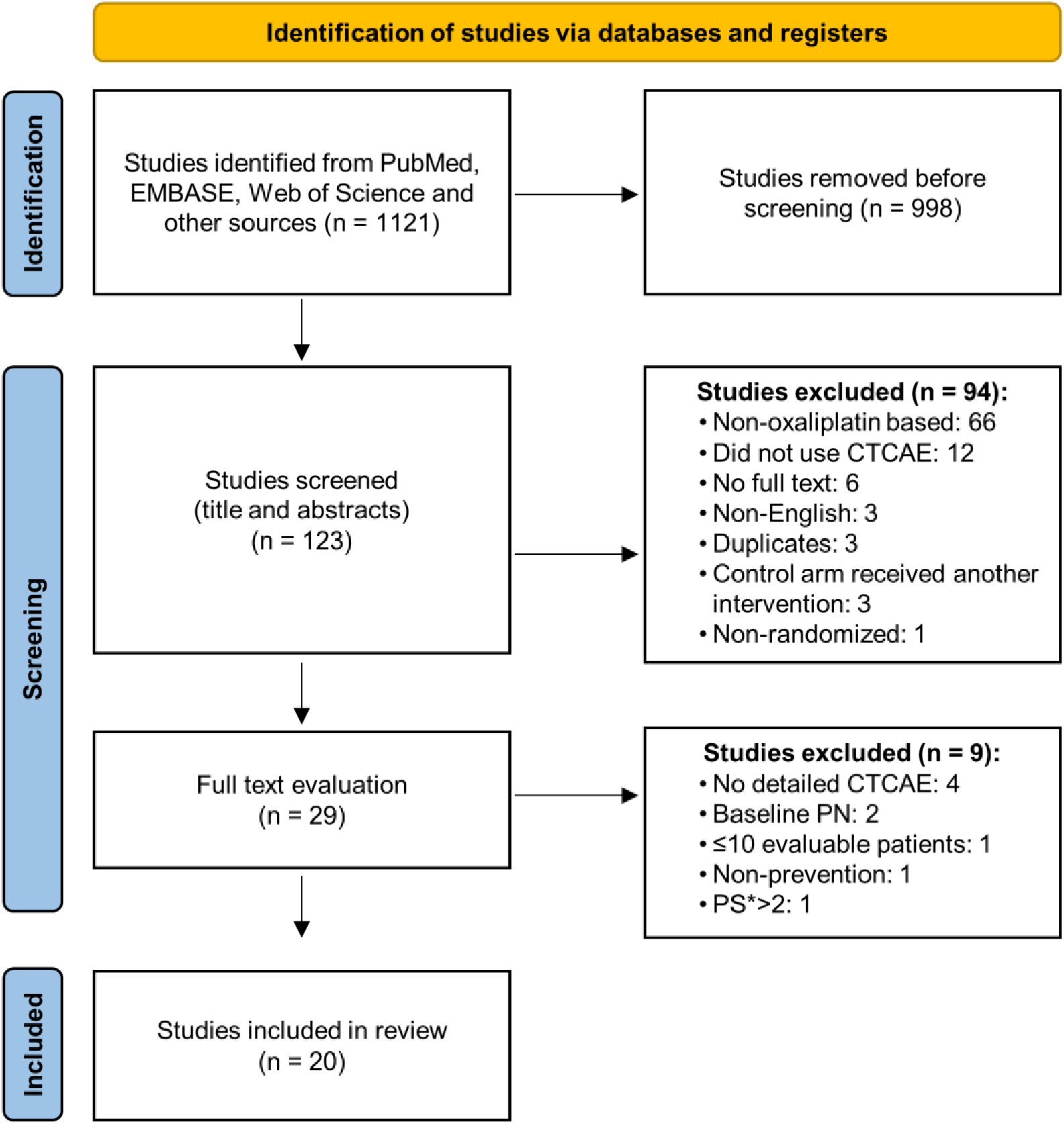
PRISMA flow diagram showing how studies were reviewed and selected. Identification, screening and exclusion criteria provided. The numbers of publications excluded at each step, and the reasons for this are indicated. CTCAE= common toxicity criteria adverse event, PN=peripheral neuropathy, PS= Performance status.

**Table 1.** Summary of included trials. The cancer type, treatment intention and the number of patients in the placebo and intervention groups is indicated for each trial. Δ included gastric cancer patients and CRC patients. M=male, F=female. PS=Performance Status. *Indicates an agent that has antioxidant scavenger properties or boosts anti-oxidative stress defences. ≠ included oesophageal cancer patients as well as CRC patient

| Author | Study Population | Regimen<br>Dose of oxaliplatin<br>Treatment schedule<br>Treatment intent | Pharmacological<br>Intervention tested<br>(N) | Control<br>(N) | Timepoints when ≥grade 2 OIPN was<br>assessed |
| --- | --- | --- | --- | --- | --- |
| Bondad et al,<br>2020<sup>28Δ</sup> | • Country: Iran. • Ethnicity: not provided. • Age (median and range): overall: 59.5 (52–65.5), intervention: 57 (52–63), control: 62.5 (53–67). • Sex: M: F: 17/15; intervention: 7/9; control: 10/6. • Cancer types: CRC (N=24), gastric (N=8); intervention: 11 CRC and 5 gastric; control: 13 CRC and 13 gastric, • Stage of disease: not provided. • PS: not provided. | XELOX<br>130 mg/m<sup>2</sup><br>Every 3 weeks<br>Adjuvant | N-Acetylcysteine*<br>(16) | Placebo<br>(16) | Baseline and at each 3-week cycle |
| Cascinu et al,<br>2002<sup>29</sup> | • Country: Italy. • Ethnicity: not provided. • Median age: intervention: 65, control: 65. • Sex: M: F: 31/21; intervention: 12/14; control: 19/7. • Cancer types: CRC. • Stages of disease: IV. • (PS: 0): intervention (N=17); control (N=20), (PS: 1): intervention (N=9); control (N=9). | FOLFOX<br>100 mg/m<sup>2</sup><br>Every 2 weeks<br>Palliative | Reduced glutathione*<br>(26) | Placebo<br>(26) | Baseline, after 4, 8 and 12 cycles |
| El-Fataty et al,<br>2018<sup>36</sup> | • Country: Egypt. • Ethnicity: not provided. • Median age: not provided. • Sex: not provided. • Cancer types: CRC. • Stages of disease: III. • PS: 0-1. | FOLFOX4<br>85 mg/m<sup>2</sup><br>Every 2 weeks<br>Adjuvant | Metformin*<br>(20) | Placebo<br>(20) | Baseline and at each 2-week cycle |
| Lin et al,<br>2006<sup>37</sup> | • Country: China. • Ethnicity: not provided. • Age (median and range): overall: 63.5 (41-78), intervention: 58 (41-75), control: 65 (43-78). • Sex: M: F: 9/5; intervention: 4/1; control: 5/4. • Cancer types: CRC. • Stages of disease: III. • (PS: 0): intervention (N=4); control (N=8), (PS: 1): intervention (N=1); control (N=1). | FOLFOX4<br>85 mg/m<sup>2</sup><br>Every 2 weeks<br>Adjuvant | N-Acetylcysteine*<br>(5) | Placebo<br>(9) | Baseline and at each 2-week cycle |
| Milla et al,<br>2009<sup>38</sup> | • Country: Italy. • Ethnicity: not provided. • Age (median and range): overall: 61 (44-75). • Sex: M: F: 18/9. • Cancer types: CRC. • Stages of disease: II & III. • PS: 0-1. | FOLFOX4<br>85 mg/m<sup>2</sup><br>Every 2 weeks<br>Adjuvant | Reduced glutathione*<br>(14) | Placebo<br>(13) | Baseline, at 5th, 9th, and 12<sup>th</sup> cycles or if the patient showed symptoms, and repeated 3 and 6 months after the last cycle |
| Yehia et al,<br>2019<sup>40</sup> | • Country: Egypt. • Ethnicity: not provided. • Mean age: overall: 45.8 ± 9.6, intervention: 45.6 ± 10.5, control: 45.9 ± 8.6. • Sex: M: F: 29/32; intervention: 16/14; control: 13/18. • Cancer types: CRC. • Stages of disease: IV. • PS: not provided. | FOLFOX6<br>85 mg/m<sup>2</sup><br>Every 2 weeks<br>Palliative | L-Carnosine*<br>(30) | Placebo<br>(31) | 3 months |
| Gobran,<br>2013<sup>22</sup> | • Country: Egypt. • Ethnicity: not provided. • Mean age: intervention: 46.5 ± 12, control: 45.9 ± 13.8. • Sex: M: F: 34/26; intervention: 16/14; control: 18/12. • Cancer types: CRC. • Stages of disease: II: intervention (N=11), control: (N=13), III: intervention (N=18), control: (N=17), IV: intervention (N=1), control: (N=0). • PS: 0-1. | FOLFOX4,<br>mFOLFOX6, FLOX<br>85 mg/m<sup>2</sup><br>Every 2 weeks<br>Adjuvant & Palliative | Ca<sup>2+</sup>/Mg<sup>2+</sup> before and<br>after Chemotherapy<br>(30) | Placebo<br>(30) | Baseline and at each 2-week cycle |
| Grothey et al,<br>2011<sup>23</sup> | • Country: USA. • Ethnicity: predominantly White (96%). • Age: <65: intervention (N=33), control: (N=33). • Sex: M: F: 54/48; intervention: 27/23; control: 27/25. • Cancer types: CRC. • Stages of disease: II & III. • PS: Not provided. | FOLFOX4,<br>mFOLFOX6 +/-<br>bevacizumab or<br>Cetuximab<br>85 mg/m<sup>2</sup><br>Every 2 weeks<br>Adjuvant | Ca<sup>2+</sup>/Mg<sup>2+</sup> before and<br>after Chemotherapy<br>(50) | Placebo<br>(52) | Baseline and at each 2-week cycle |
| Ishibashi et al, 2010<sup>24</sup> | Country: Japan.<br>Ethnicity: not provided.<br>Age (median and range): intervention: 63 (32–74), control: 64 (35–73).<br>Sex: M: F: 16/17; intervention: 9/8; control: 7/9.<br>Cancer types: CRC.<br>Stages of disease: IV.<br>PS: 0-2. | mFOLFOX6<br>85 mg/m<sup>2</sup><br>Every 2 weeks<br>Adjuvant & Palliative | Ca<sup>2+</sup>/Mg<sup>2+</sup> before and after Chemotherapy (17) | Placebo (16) | Baseline and at each 2-week cycle |
| Loprinzi et al, 2014<sup>25</sup> | Country: USA.<br>Ethnicity: White: intervention 1 (N=96), intervention 2 (N=99), control (N=105), Black or African American: intervention 1 (N=16), intervention 2 (N=15), control (N=10), Asian: intervention 1 (N=4), intervention 2 (N=1), control (N=2), American Indian or Alaska Native: intervention 1 (N=0), intervention 2 (N=1), control (N=1).<br>Median age: overall: 56, intervention 1: 57, intervention 2: 57, control: 56.<br>Sex: M: F: 169/184, intervention 1 (56/62), intervention 2 (56/60), control: (57/62).<br>Cancer types: CRC.<br>Stages of disease: II: intervention 1 (N=22), intervention 2 (N=22), control (N=22), III: intervention 1 (N=89), intervention 2 (N=88), control (N=88), IV: intervention 1 (N=7), intervention 2 (N=6), control (N=9).<br>PS: Not provided. | FOLFOX4 or mFOLFOX6<br>85 mg/m<sup>2</sup><br>Every 2 weeks<br>Adjuvant | Intervention 1: Ca<sup>2+</sup>/Mg<sup>2+</sup> before and after chemotherapy (118)<br>Intervention 2: Ca<sup>2+</sup>/Mg<sup>2+</sup> before and placebo after chemotherapy (116) | Placebo (119) | Baseline and at each 2-week cycle |
| Kono et al, 2013<sup>30</sup> | Country: Japan.<br>Ethnicity: not provided.<br>Age (median and range): intervention: 67 (40–88), control: 61 (36–82).<br>Sex: M: F: 48/41; intervention: 23/21; control: 25/20.<br>Cancer types: CRC.<br>Stages of disease: IV: intervention (N=36); control (N=35), non-metastatic: intervention (N=8); control (N=10).<br>(PS: 0): intervention (N=40); control (N=44), (PS:1): intervention (N=4); control (N=1). | FOLFOX4 or mFOLFOX6<br>85mg/m<sup>2</sup><br>Every 2 weeks<br>Adjuvant & palliative | Goshajinkigan (44) | Placebo (45) | Baseline, every 2 weeks until the 8th cycle, and every 4 weeks thereafter until the 26th week |
| Oki et al, 2015<sup>31</sup> | Country: Japan.<br>Ethnicity: not provided.<br>Median age: intervention: 62.4 ± 10.6, control: 60.4 ± 11.5.<br>Sex: M: F: 99/83 intervention: 48/41; control: 51/42.<br>Cancer types: CRC.<br>Stages of disease: III. (PS: 0): intervention (N=85); control (N=92), (PS:1): intervention (N=4); control (N=1). | mFOLFOX6<br>85mg/m<sup>2</sup><br>Adjuvant | Goshajinkigan (89) | Placebo (93) | Baseline and at each 2-week cycle |
| Aghili et al, 2023<sup>32#</sup> | Country: Iran.<br>Ethnicity: not provided.<br>Mean age: intervention: 53 ± 12.4, control: 55 ± 11.6.<br>Sex: M: F: 24/8; intervention: 12/5; control: 12/3.<br>Cancer types: CRC: intervention (N=17); control (N=13), Oesophageal cancer: intervention (N=0); control (N=2).<br>Stages of disease: IV: intervention (N=2); control (N=4), non-metastatic: intervention (N=15); control (N=11).<br>PS: 0-1. | CAPOX<br>130 mg/m<sup>2</sup><br>Every 3 weeks<br>Adjuvant & palliative | Duloxetine (17) | Placebo (15) | 1 day before and within 1 week after each cycle |
| Kobayashi et al, 2020<sup>41</sup> | Country: Japan.<br>Ethnicity: not provided.<br>Mean age: intervention: 58.1 ± 2.7, control: 67.5 ± 1.7.<br>Sex: M: F: 11/17; intervention: 8/6; control: 3/11.<br>Cancer types: CRC.<br>Stages of disease: II: intervention (N=0); control (N=3), III: intervention (N=9); control (N=7), IV: intervention (N=5); control (N=4).<br>PS: 0-1. | mFOLFOX6<br>85mg/m<sup>2</sup><br>Adjuvant & palliative | Cystine and theanine (14) | Placebo (14) | Baseline and at each 2-week cycle |
| Kotaka et al, 2020<sup>33</sup> | Country: Japan.<br>Ethnicity: not provided.<br>Age (median and range): intervention 1: 68.0 (38–78), intervention 2: 66.0 (32–79), control: 68.0 (45–79).<br>Sex: M: F: 39/40, intervention 1 12/15, intervention 2 11/13, control: 16/12.<br>Cancer types: CRC.<br>Stages of disease: II: intervention 1 (N=3), intervention 2 (N=6), control: (N=6), IIIa: intervention 1 (N=17), intervention 2 (N=15), control: (N=16), IIIb: intervention 1 (N=7), intervention 2 (N=3), control: (N=6).<br>(PS:0): intervention 1 (N=27), intervention 2 (N=24), control (N=25), (PS:1): intervention 1 (N=0), intervention 2 (N=0), control: (N=3). | mFOLFOX6<br>85mg/m<sup>2</sup><br>Every 2 weeks<br>Adjuvant | Intervention1: 1-day ART-123 (27)<br>Intervention2: 3-day ART-123 (24) | Placebo (28) | Every day from day 1 to day 3 of each cycle and on days 15 and 43 of cycle 12 |
| Lee et al, 2023<sup>39</sup> | Country: US and Canada.<br>Ethnicity: White (79.3%).<br>Mean age: 60.9, Sex: M: F: 1098/1352.<br>Cancer types: CRC.<br>Stages of disease: III.<br>PS: 0: N=1741, 1-2: 709. | FOLFOX<br>85mg/m<sup>2</sup><br>Every 2 weeks<br>Adjuvant | Celecoxib<br>(1228) | Placebo<br>(1222) | Every 3 months for 3 years and subsequently every 6 months for 6 years after random assignment or until disease recurrence. |
| Liu et al, 2013<sup>34</sup> | Country: China.<br>Ethnicity: not provided.<br>Mean age: 52.5, ≥50: intervention: 10, control: 7, <50: Intervention: 50, control: 53.<br>Sex: M: F: 83/37; intervention: 43/17; control: 40/20.<br>Cancer types: CRC.<br>Stages of disease: IV.<br>(PS: 0): intervention (N=34), control: (N=29), (PS:1-2): intervention (N=26), control: (N=31). | FOLFOX4<br>85mg/m<sup>2</sup><br>Every 2 weeks<br>Palliative | Guilongtongluoang<br>(60) | Placebo<br>(60) | Baseline and every two cycles for 6 cycles |
| Motoo et al, 2020<sup>42</sup> | Country: Japan.<br>Ethnicity: not provided.<br>Age (median and range): intervention: 62 (35-79), control: 68 (53-79).<br>Sex: M: F: 31/21; intervention: 16/10; control: 15/11.<br>Cancer types: CRC.<br>Stages of disease: III.<br>(PS: 0): intervention (N=20), control: (N=20), (PS:1): intervention (N=6), control: (N=6). | CAPOX<br>130 mg/m<sup>2</sup><br>Every 3 weeks<br>Adjuvant | Ninjin'yoeito<br>(26) | Placebo<br>(26) | Baseline and at each 3-week cycle |
| Wang et al, 2020<sup>35</sup> | Country: China.<br>Ethnicity: not provided.<br>Median age: intervention: 52, control: 55.5.<br>Sex: M: F: 111/85; intervention: 53/45; control: 58/40.<br>Cancer types: CRC.<br>Stages of disease: II: intervention (N=26); control (N=38), III: intervention (N=72); control (N=60).<br>PS: Not provided. | mFOLFOX6<br>85mg/m<sup>2</sup><br>Every 2 weeks<br>Adjuvant | Monosialotetraexosyl-ganglioside<br>(96) | Placebo<br>(96) | Baseline and at each 2-week cycle |
| Zhang et al, 2012<sup>43</sup> | Country: China.<br>Ethnicity: not provided.<br>Median age: intervention: 55.1, control: 57.3.<br>Sex: M: F: 52/27; intervention: 25/13; control: 27/14.<br>Cancer types: CRC.<br>Stages of disease: II: intervention (N=13); control (N=18), III: intervention (N=25); control (N=23).<br>PS: Not provided. | CAPOX<br>130 mg/m<sup>2</sup><br>Every 3 weeks<br>Adjuvant | Neurotropin<br>(38) | Placebo<br>(41) | Every week by an Investigator and every 3 weeks by two clinicians |

The studies were conducted in multiple countries, six in Japan,^24,30,31,33,41,42^ four from China,^34,35,37,43^ three from Egypt,^22,36,40^ two from each of USA,^23,25^ Iran^28,32^ and Italy,^29,38^ and one from each of USA and Canada^39^ (Table 1). A total of 1989 patients were given interventional treatments and 1972 control patients were included. 1917 males versus 2008 females were enrolled into the trials and where ethnicity was reported, the majority were White. In most studies the enrolled patients had stage III or IV CRC. The doses of oxaliplatin used to treat patients were 85,^22–25,30,31,33–41^ 100^29^ or 130^28,32,42,43^ mg/m^2^ every 2-3 weeks. 14 different pharmacological interventions were investigated: N-acetylcysteine,^28,37^ glutathione,^29,38^ metformin,^36^ L-carnosine,^40^ Ca^2+^/Mg^2+^ infusions,^22,23,24,25^ goshajinkigan,^30,31^ monosialotetrahexosylganglioside,^35^ cystine and theanine,^41^ recombinant thrombomodulin (ART-123),^33^ guilongtongluofang,^34^ ninjin’yoeito,^42^ neurotropin,^43^ duloxetine^32^ and celecoxib.^39^ Included in two separate meta-analysis were firstly, N-acetylcysteine, glutathione, metformin and L-carnosine, grouped because each is able to support anti-oxidative stress responses in cells either directly via scavenger activity, by inducing antioxidant transcription or by supporting mitochondrial health and function; and secondly, Ca^2+^/Mg^2+^ infusions. OIPN was assessed every 2 weeks in most studies (12/20 studies, Table 1), (range: weekly to every 3 months). Supplementary Table 3 includes details of 17 agents that we excluded from the review because trials did not measure OIPN using the CTCAE scale.

### Risk of bias

The risk of bias analysis showed that 35% of studies had potential high risk of bias with 45% of studies showing low risks of bias (Figure 2A and B). Only domain 2, which assessed risk of bias due to deviations from the intended interventions, show that all studies possessed a low risk of bias. However, all other domains showed potential for high risk of bias in 10-35% of the studies.

**Figure 2.**
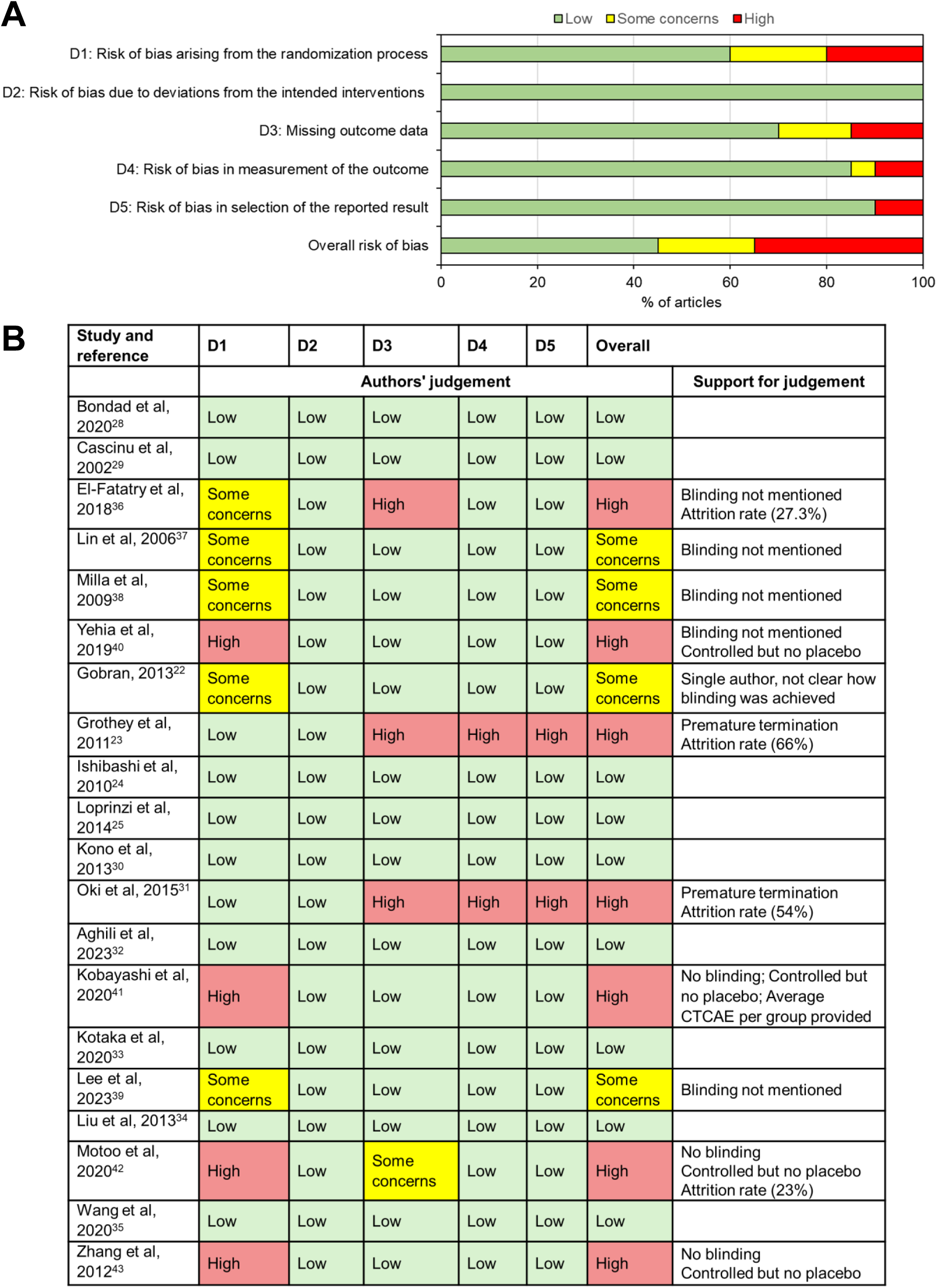
Risk of bias analysis. (**A**) Summary chart to show the proportions of studies with risk of bias in each domain assessed. (**B**) Risk of bias in individual studies with explanations for the authors’ judgement.

### Interventions evaluated by single studies

Eight interventions were investigated by only a single study: monosialotetrahexosylganglioside,^35^ ART-123,^33^ duloxetine^32^ and celecoxib.^39^ None showed a significant reduction in grade ≥2 OIPN in clinical trials of 192, 51, 32 and 2,450 patients with CRC, respectively. In the celecoxib trial,^39^ patients were randomised to 6 or 12 weeks of oxaliplatin ± 3 years of celecoxib treatment. 1,225 patients received celecoxib and there was no difference in incidence of grade ≥2 OIPN in this group compared to the placebo-controlled group either during or after completion of treatment with oxaliplatin. There was also no difference in severity of OIPN measured 12 months after completion of treatment with oxaliplatin using the Functional Assessment of Cancer Therapy/Gynecologic Oncology Group-Neurotoxicity-13 (FACT/GOG-NTX-13). Thanks to this well powered trial there is strong evidence against the use of celecoxib to treat OIPN in CRC patients. The other single trials evaluating agents were too small to draw definitive conclusions.

Oral administration of cystine and theanine,^41^ guilongtongluofang,^34^ ninjin’yoeito^42^ and neurotropin^43^ showed statistically significant association with reduced grade ≥2 OIPN (results summarised in Supplementary Table 4). The sample sizes employed by these clinical trials ranged from 28-120. All these interventions need evaluating in future independent trials.

### Goshajinkigan

Goshajinkigan (TJ-107) is a traditional Japanese herbal remedy used for pain relief. Previous meta-analyses of studies investigating whether goshajinkigan is effective in reducing taxane-induced PN concluded there was no significant effect.^44^ We identified 2 fully-blinded, placebo-controlled trials that evaluated whether goshajinkigan could ameliorate OIPN.^30,31^ The first, Kono *et al*, 2013^30^ enrolled 93 patients, of which 89 could be included in the analysis. A non significant reduction in grade ≥2 and grade ≥3 OIPN at cycle 8 was observed in the arm randomised to receive goshajinkigan (39% *vs.* 51% in placebo arm and 7% *vs.* 13%, respectively). The authors also investigated tumour response in 27 patients receiving goshajinkigan and 23 receiving placebo and observed a non significant improvement in response in the patients receiving goshajinkigan. The second study, Oki *et al*, 2015^31^ performed an interim analysis of 142 patients and identified a significantly higher incidence of grade ≥2 OIPN in the arm receiving goshajinkigan compared to placebo (50.6% *vs.* 39%), and a significantly shorter time to grade ≥2 OIPN in the goshajinkigan arm (RR=1.908, 95% CI: [1.81-3.083], p=0.007). The interim analysis was performed using assessments of OIPN up until the time when 50% of the planned recruitment had been achieved. All patients had received at least one cycle of oxaliplatin but a breakdown of the number of cycles patients included in the interim analysis had received was not provided. Since it had been determined that goshajinkigan could not significantly outperform placebo even if the study was allowed to continue, it was terminated early. Collectively, these two studies do not support goshajinkigan being used to treat grade ≥2 OIPN.

### Interventions evaluated by three or more studies

Two interventions trialled in three or more studies were analysed by random-effects meta-analysis. Multiple therapies able to increase anti-oxidative stress responses or defences in cells were evaluated together.^28,29,36,37,38,40^

### Interventions targeting oxidative stress

We analysed 6 trials ^28,29,36,37,38,40^ investigating 4 drugs that have anti-oxidative stress effects: N-acetylcysteine,^28,37^ glutathione,^29, 38^ L-carnosine^40^ and metformin.^36^ N-acetylcysteine is a precursor of glutathione, a key antioxidant molecule and administration of each raises cellular glutathione levels. Metformin and L-carnosine induce antioxidant defences indirectly through effects on mitochondria and transcription and L-carnosine also has direct free radical scavenger effects and chelates Zn^2+^.^45,46^ Interestingly each of the studies investigating a drug with anti-oxidative stress effects found significant reduction in CTCAE grade ≥2 OIPN in their intervention arms. Five of the studies assessed OIPN at the end of 8-12 cycles (6 months) therapy^28,29,36,38,37^ and three assessed OIPN after 4-6 cycles (2-3 months of treatment)^29,36,40^ so two separate meta-analyses were performed (Figures 3 and 4). Administration of anti-oxidative stress agents was not associated with a statistically significant reduced risk of grade ≥2 OIPN after 2-3 months of treatment (Figure 3; OR: 0.19, 95% CI: [0.02 -1.60], p=0.013) but, when assessed after 8-12 cycles (6 months of treatment), a statistically significant reduction of large effect was observed (Figure 4; OR: 0.04, 95% CI: [0.02-0.12], p<0.00001). High heterogeneity was observed in the analysis of 2-3 months of treatment outcomes (I^2^=66%) but no heterogeneity was observed in the analysis of 6 months of treatment outcomes (I^2^=0%).

**Figure 3.**
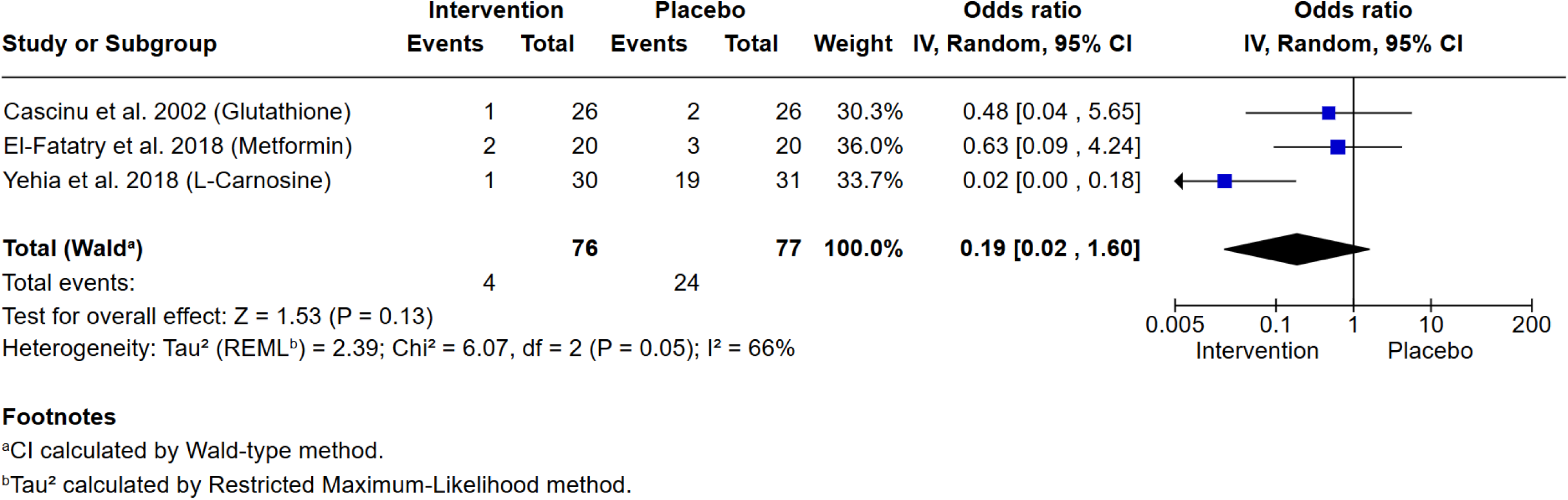
Treatment with anti-oxidative stress drugs is not associated with a significant reduction in CTCAE grade ≥2 peripheral neuropathy after 4-6 cycles of oxaliplatin-based chemotherapy. Meta-analysis of 3 studies shows no significant reduction in incidence of neuropathy *vs.* placebo (P = 0.13). The intervention tested in each trial is indicated in parentheses.

**Figure 4.**
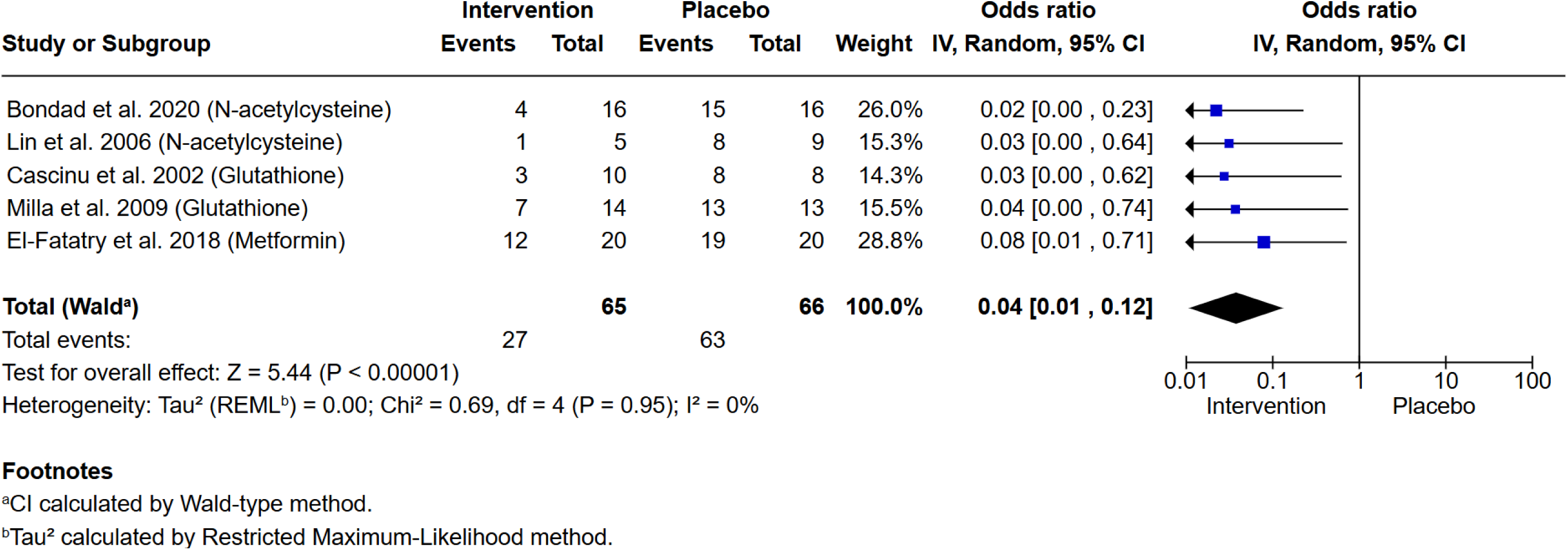
Treatment with anti-oxidative stress drugs is associated with a significant reduction in CTCAE grade ≥2 peripheral neuropathy after 8-12 cycles of oxaliplatin-based chemotherapy for colorectal cancer,. Meta-analysis of 5 studies shows reduced incidence of neuropathy *vs.* placebo (P<0.00001). The intervention tested in each trial is indicated in parentheses.

The study by Yehia *et al*, 2019^40^ that tested L-carnosine was not placebo controlled – a control arm was included, but no blinding. A sensitivity analysis was performed to check whether a similar effect size was seen when assessing OIPN after 2-3 months of oxaliplatin if this study was excluded (Supplementary Figure 1). Glutathione and metformin intervention arms showed no significant difference in grade 2 OIPN compared to the control (OR: 0.57, 95% CI: [0.13-2.57], p=0.46). Only 2 of the studies included in the analysis of OIPN at 8-12 cycles were double-blind, placebo-controlled trials. Supplementary Figure 2 shows the meta-analysis of Bondad *et al*, 2020^28^ and Cascinu *et al*. 2002^29^ which tested N-acetylcysteine and glutathione, respectively, but with the 3 non-blinded trials excluded. Very similar overall measures of effect are seen compared to the 5-study meta-analysis presented in Figure 4, (OR: 0.02, 95% CI: [0.00-0.15], p<0.0001).

### Ca^2+/^Mg^2+^ infusions

Three double-blinded, placebo-controlled trials ^22,23,25^ investigated patients randomised to receive either an infusion of Ca^2+^/Mg^2+^ pre- and post-oxaliplatin treatment (198 patients in total) or placebo (200 patients). One rationale for Ca^2+^/Mg^2+^ infusions was that acute PN may be due to the disruption of ion levels that affects neuronal function and homeostasis.^47^ Meta-analysis of the three trials revealed no association between administration of Ca^2+^/Mg^2+^ infusions pre- and post-oxaliplatin treatment and risk of CTCAE grade ≥2 PN, as assessed after 6 months of oxaliplatin-based chemotherapy (Figure 5; OR: 0.57, 95% CI: [0.29-1.11], p*=*0.10). This is in keeping with the results of a meta-analysis by Peng *et al*, 2022.^13^ The Loprinzi *et al*, 2014^25^ trial also included an arm where patients received Ca^2+^/Mg^2+^ before oxaliplatin and placebo after. It also failed to show any significant reduction of OIPN (p*=*0.3383). The risk of bias analysis (Figure 2A and B) identified high risk of bias for the Grothey at al, 2011 study^23^ and noted a high attrition rate, as well as premature termination following interim results from the Combined Oxaliplatin Neurotoxicity Prevention Trial (CONcepT) which suggested Ca^2+^/Mg^2+^ infusions affected the response to chemotherapy.^48^

**Figure 5.**
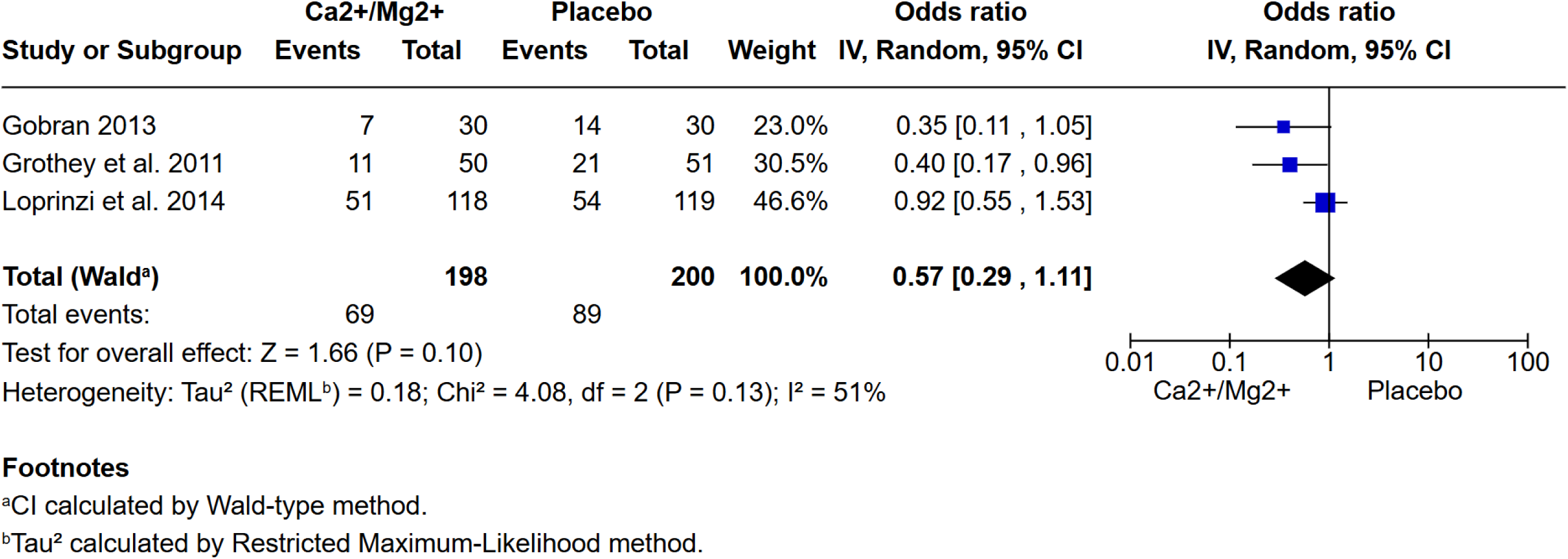
The cumulative incidence of CTCAE grade ≥2 peripheral neuropathy after 6 months of oxaliplatin-based chemotherapy is not reduced by administration of Ca^2+^/Mg^2+^ infusions before and after chemotherapy. Meta-analysis of 3 studies shows no significant difference in the incidence of OIPN *vs.* placebo (P*=* 0.10).

## Discussion

In this systematic review we set out to ask whether there is any evidence that pharmacological agents or traditional herbal medicines are able to reduce CTCAE grade ≥2 OIPN in CRC patients. We applied strict inclusion criteria to evaluate the evidence from current RCTs that have tested 14 pharmacological interventions and traditional herbal medicines. Two interventions were subsequently analysed by meta-analysis: first, a group of drugs that all have anti-oxidative stress effects in cells; and second, Ca^2+^/Mg^2+^ infusions. We found that only anti-oxidative stress drugs were associated with a lower risk of grade ≥2 OIPN when assessed at the end of the treatment (8-12 cycles). There was no significant effect on grade ≥2 OIPN mid-treatment, regardless of whether we included or removed the study by Yehia *et al*.^40^ which was not placebo-controlled or blinded.

Consistent with previous reviews,^11,12,13^ no protective effect of Ca^2+^/Mg^2+^ infusions on CTCAE grade ≥2 OIPN was detected. Similarly, there was no evidence of a protective effect of goshajinkigan (TJ-107) in two independent studies.^30,31^ All other pharmacological interventions were investigated in only single studies and, with the exception of celecoxib which was investigated in a very large trial with a long follow-up,^39^ all other agents will need further need evaluation. Future studies will also need to consider whether treatment with pharmacological agents to reduce OIPN affects the efficacy of the oxaliplatin chemotherapy since most of the RCT reviewed here were too small, or followed up patients for too short a time, to draw conclusions.

A recently published meta-analysis also studied interventions aiming to reduce OIPN.^13^ Supplementary Table 4 summarises the details of the 43 studies they included and the reason why each study was, or was not, included in our study. We used stricter inclusion critera in our study. First, we only analysed data from studies that had assessed OIPN using the CTCAE criteria. Second, we only combined datasets when OIPN was assessed at a similar timepoint in the chemotherapy regime because the severity of PN does change over the course of treatment with oxaliplatin.^6^ A high proportion of patients (>85%) experience acute neurotoxicity but this often resolves 4-6 weeks into treatment.^49^ Third, we excluded some studies where we could not access a full text version in English. Fourth, we excluded studies where some patients had baseline PN. Fifth, we excluded studies with fewer than 10 participants available for the final analysis. Twenty studies met our inclusion criteria but the majority of the RCTs recruited small numbers of participants to each arm of their study, with only five studies randomising more than 50 patients to each arm.^39,25,31,34,35^ The risk of bias analysis highlights that 7 of the included studies had a high risk of bias in at least one of the assessed categories.^36,40,23,31,41,42,43^

### The eJect of anti-oxidative stress treatments on OIPN

While the benefical effect of administrating anti-oxidative stress drugs alongside oxaliplatin was clear from the meta-analysis that we performed, the drugs do not all act by the same mechanism.

1. *Glutathione and N-acetylcysteine.* N-acetylcysteine is a precursor of glutathione and administration leads to an increase in glutatione levels in cells.^29^ Reduced glutathione is a principal scavenger of free radicals,^50^ hence administration of either N-acetylcysteine or glutathione directly can be considered to be acting via the same mechanism: increasing scavenger levels that reduce reactive species.
2. *L-carnosine* (β-alanyl-L-histidine) is a di-peptide synthesised predominantly in muscles and in glia with multiple antioxidant and anti-inflammatory roles.^51^ L-carnosine acts directly as a scavenger of reactive oxygen and nitrogen species but it also has more indirect effects on cellular defences by inducing Nrf2-dependent transcription. Nrf2 is a key transcription factor that coordinates a large network of anti-oxidative stress transcription *via* the antioxidant response element (ARE).^52^ L-carnosine is also able to form a complex with Zn^2+^ to generate polaprezinc which has anti-inflammatory and antioxidant effects, thought in part to be due to upregulation of heme oxygenase.^53^ Importantly when considering inteventions to treat OIPN, L-carnosine is orally bioavailable and brain-penetrant. Having said that, the oral bioavailability of L-carnosine is reduced by the action of serum carnosinases and testing of analogues such as carnosinol that are more resistant to carnosinase activity, ^54^ may be preferable in future trials.
3. *Metformin* (1,1-dimethylbiguanide) is a first-line treatment for type II diabetes and is also commonly prescribed for other glucose-resistance disorders, such as polycystic ovary syndrome.^55^ It has also been tested in numerous pre-clinical models of acute neurological injury and chronic neurological disease, including as a method of reducing peripheral neuropathy.^36^ Metformin reduces gluconeogenesis in the liver and increases insulin uptake by cells, although the key biochemical pathways are debated. Metformin can inhibit mitochondrial complex I activity.^56^ This is likely to be especially relevant to OIPN since oxaliplatin and other platinum drugs have been shown in numerous studies to increase production of reactive oxygen species by mitochondria.^8^ Inhibition of complex I will also activate AMP-activated protein kinase (AMPK) and subsequently induce autophagy, which has been shown to be neuroprotective in numerous models of neurological disease. Metformin can also activate AMPK more directly via Liver kinase B1 (LKB1).^57^ Importantly, metformin alters the redox balance of cells by increasing levels of NADPH. Given, that NADPH is required to maintain the cellular pool of reduced glutathione, metformin treatment leads to a general increase in cellular oxidative stress defences.

Given that oxaliplatin treatment results in mitochondrial dysfunction and excess production of ROS, it seems probable, therefore, that anti-oxidative stress compounds protect against PN by scavenging excess ROS and/or by inducing Nrf2-based anti-oxidant transcription. If true, it would be interesting to test targetted forms of antioxidants that concentrate in mitochondria e.g. mitoQ, as these may be more effective at lower doses.

### Treatment of OPIN with anti-oxidative stress drugs

Previous studies have uncovered potentially protective effects of anti-oxidative stress agents against CIPN^58,59^. They included patients with different cancer types, treated with various different chemotherapy agents and scored PN using various different methods. This study is, to our knowledge, the first to systematically review potentially protective treaments for severe PN in patients with the same cancer type, treated with the same chemotherapy drug and where the same method of scoring PN was used in each trial.

None of N-acetylcysteine, glutathione, L-carnosine or metformin are currently recommended for treating OPIN in the current ASCO guidelines^12^ as there is a lack of evidence from a large clinical trial that any provide a benefit. The large effect size of anti-oxidative stress compounds revealed in our meta-analysis, coupled with the lack of heterogeneity in the meta-analysis when OIPN was assessed at 6 months of treatment, supports a larger study to confirm this protective effect. This study must also address potential changes to the efficacy of oxaliplatin. These anti-oxidative stress compounds are cheap and well-tolerated. A successful trial would open options for a simple way to treat OPIN simply with large potential benefit for CRC patients.

### Limitations of the study

The studies included in the meta-analysis were all small and each had a high rate of grade ≥2 OIPN in the placebo arms (88-100%). It is possible that these interventions will not prove effective at reducing incidence of grade ≥2 OIPN if the incidence in the placebo arm is lower. We only considered trials using clinician-scored CTCAE criteria to score OIPN to facilitate comparability across studies. However, we accept that it is a subjective measure and will vary between the scorers and this may not be the optimal measure of OIPN symptom severity; patient reported outcomes may be preferable, particularly if combined with CTCAE scores.^60^

Despite strict inclusion criteria there were potential sources of bias identified in 4 out of the 6 studies that investigated anti-oxidative stress agents.^36,37,38,40^ Blinding was not clearly described for four of the studies. ^36,37,38,40^ However, when unblinded trials were excluded, the two remaining studies ^28,29^ also showed clear evidence of a significant reduction in grade ≥2 OIPN (*P<*0.0001) in the intervention arm in 50 patients (Supplementary figure 2).

Further validation through a large-scale, high quality RCT is essential to establish a definitive beneficial effect and to identify which class of antioxidants may be most effective and best tolerated. There remains an urgent need for novel interventions to reduce peripheral neuropathy and enhance the quality of life for patients receiving oxaliplatin-based therapies.

## Data Availability

All data produced in the present work are contained in the manuscript

## Acknowledgements

This work was supported by Cancer Research UK (Ref SEBCATP-2023/100001). The authors declare no conflict of interest.

The research was carried out at the National Institute for Health and Care Research (NIHR) Birmingham Biomedical Research Centre (BRC).

## References

1. Maihöfner, C., Diel, I., Tesch, H., Quandel, T. & Baron, R. Chemotherapy-induced peripheral neuropathy (CIPN): current therapies and topical treatment option with high-concentration capsaicin. Supportive Care in Cancer 29, 4223 (2021).

2. Zajaczkowską, R. et al. Mechanisms of chemotherapy-induced peripheral neuropathy. Int J Mol Sci 20, (2019).

3. Rokhsareh, S., Haghighi, S. & Tavakoli-Ardakani, M. Evaluating the effects of duloxetine on prophylaxis of oxaliplatin-induced peripheral neuropathy in patients with gastrointestinal cancer: A randomized double-blind placebo controlled clinical trial. Journal of Oncology Pharmacy Practice 29, 60–65 (2023).

4. Argyriou, A. A. et al. Clinical pattern and associations of oxaliplatin acute neurotoxicity: a prospective study in 170 patients with colorectal cancer. Cancer 119, 438–444 (2013).

5. Maestri, A. et al. A pilot study on the effect of acetyl-L-carnitine in paclitaxel- and cisplatin-induced peripheral neuropathy. Tumori 91, 135–138 (2005).

6. Teng, C., Cohen, J., Egger, S., Blinman, P. L. & Vardy, J. L. Systematic review of long-term chemotherapy-induced peripheral neuropathy (CIPN) following adjuvant oxaliplatin for colorectal cancer. Supportive Care in Cancer 30, 33–47 (2022).

7. Sałat, K. Chemotherapy-induced peripheral neuropathy—part 2: focus on the prevention of oxaliplatin-induced neurotoxicity. Pharmacological Reports 72, 508–527 (2020).

8. Colvin, L. A. Chemotherapy-induced peripheral neuropathy: Where are we now? Pain 160, S1–S10 (2019).

9. NHS Digital. Cancer Registrations Statistics, England 2021-First release, counts only. 1–23 (2023).

10. Jordan, B. et al. Systemic anticancer therapy-induced peripheral and central neurotoxicity: ESMO–EONS–EANO Clinical Practice Guidelines for diagnosis, prevention, treatment and follow-up. Annals of Oncology 31, 1306–1319 (2020).

11. Hershman, D. L. et al. Prevention and management of chemotherapy-induced peripheral neuropathy in survivors of adult cancers: American society of clinical oncology clinical practice guideline. Journal of Clinical Oncology 32, 1941–1967 (2014).

12. Loprinzi, C. L. et al. Prevention and management of chemotherapy-induced peripheral neuropathy in survivors of adult cancers: ASCO guideline update. Journal of Clinical Oncology 38, 3325–3348 (2020).

13. Peng, S. et al. Prevention of Oxaliplatin-Induced Peripheral Neuropathy: A Systematic Review and Meta-Analysis. Front Oncol 12, 731223 (2022).

14. Glimelius, B. et al. Persistent prevention of oxaliplatin-induced peripheral neuropathy using calmangafodipir (PledOx®): a placebo-controlled randomised phase II study (PLIANT). Acta Oncol (Madr) 57, 393–402 (2018).

15. Wang, W.-S. et al. Oral Glutamine Is Effective for Preventing Oxaliplatin-Induced Neuropathy in Colorectal Cancer Patients. Oncologist 12, 312–319 (2007).

16. de Andrade, D. C. et al. Pregabalin for the Prevention of Oxaliplatin-Induced Painful Neuropathy: A Randomized, Double-Blind Trial. Oncologist 22, 1154–e105 (2017).

17. Bruna, J. et al. Efficacy of a Novel Sigma-1 Receptor Antagonist for Oxaliplatin-Induced Neuropathy: A Randomized, Double-Blind, Placebo-Controlled Phase IIa Clinical Trial. Neurotherapeutics 15, 178–189 (2018).

18. Vitale, M. G. et al. Zeoxanmulti trial: A Randomized, Double-Blinded, Placebo-Controlled Trial of Oral PMA-zeolite to prevent Chemotherapy-Induced Side Effects, in particular, Peripheral Neuropathy. Molecules 25, (2020).

19. Durand, J. P. et al. Efficacy of venlafaxine for the prevention and relief of oxaliplatin-induced acute neurotoxicity: results of EFFOX, a randomized, double-blind, placebo-controlled phase III trial. Ann Oncol 23, 200–205 (2012).

20. Zhang, X. et al. Prevention of oxaliplatin-related neurotoxicity by ω-3 PUFAs: A double-blind randomized study of patients receiving oxaliplatin combined with capecitabine for colon cancer. Medicine (United States) 99, E23564 (2020).

21. Chay, W. Y. et al. Use of calcium and magnesium infusions in prevention of oxaliplatin induced sensory neuropathy. Asia Pac J Clin Oncol 6, 270–277 (2010).

22. Gobran, N. S. Role of calcium and magnesium infusion in prevention of oxaliplatin neurotoxicity. A phase III trial. Chinese-German Journal of Clinical Oncology 12, 232–236 (2013).

23. Grothey, A. et al. Intravenous calcium and magnesium for oxaliplatin-induced sensory neurotoxicity in adjuvant colon cancer: NCCTG N04C7. Journal of Clinical Oncology 29, 421–427 (2011).

24. Ishibashi, K., Okada, N., Miyazaki, T., Sano, M. & Ishida, H. Effect of calcium and magnesium on neurotoxicity and blood platinum concentrations in patients receiving mFOLFOX6 therapy: a prospective randomized study. Int J Clin Oncol 15, 82–87 (2010).

25. Loprinzi, C. L. et al. Phase III randomized, placebo-controlled, double-blind study of intravenous calcium and magnesium to prevent oxaliplatin-induced sensory neurotoxicity (N08CB/Alliance). J Clin Oncol 32, 997–1005 (2014).

26. RevMan: Systematic review and meta-analysis tool for researchers worldwide | Cochrane RevMan. https://revman.cochrane.org/info.

27. Sterne, J. A. C. et al. RoB 2: a revised tool for assessing risk of bias in randomised trials. BMJ 366, (2019).

28. Bondad, N., Boostani, R., Barri, A., Elyasi, S. & Allahyari, A. Protective effect of N-acetylcysteine on oxaliplatin-induced neurotoxicity in patients with colorectal and gastric cancers: A randomized, double blind, placebo-controlled, clinical trial. Journal of Oncology Pharmacy Practice 26, 1575–1582 (2020).

29. Cascinu, S. et al. Neuroprotective effect of reduced glutathione on oxaliplatin-based chemotherapy in advanced colorectal cancer: A randomized, double-blind, placebo-controlled trial. Journal of Clinical Oncology 20, 3478–3483 (2002).

30. Kono, T. et al. Goshajinkigan oxaliplatin neurotoxicity evaluation (GONE): A phase 2, multicenter, randomized, double-blind, placebo-controlled trial of goshajinkigan to prevent oxaliplatin-induced neuropathy. Cancer Chemother Pharmacol 72, 1283–1290 (2013).

31. Oki, E. et al. Preventive effect of Goshajinkigan on peripheral neurotoxicity of FOLFOX therapy (GENIUS trial): a placebo-controlled, double-blind, randomized phase III study. Int J Clin Oncol 20, 767–775 (2015).

32. Aghili, M. et al. Duloxetine for the Prevention of Oxaliplatin Induced Peripheral Neuropathy: A Randomized, Placebo-Controlled, Double-blind Clinical Trial. J Gastrointest Cancer 54, 467–474 (2023).

33. Kotaka, M. et al. A placebo-controlled, double-blind, randomized study of recombinant thrombomodulin (ART-123) to prevent oxaliplatin-induced peripheral neuropathy. Cancer Chemother Pharmacol 86, 607–618 (2020).

34. Liu, Y. et al. Clinical study on the prevention of oxaliplatin-induced neurotoxicity with guilongtongluofang: results of a randomized, double-blind, placebo-controlled trial. Evid Based Complement Alternat Med 2013, (2013).

35. Wang, D. shen et al. Phase III randomized, placebo-controlled, double-blind study of monosialotetrahexosylganglioside for the prevention of oxaliplatin-induced peripheral neurotoxicity in stage II/III colorectal cancer. Cancer Med 9, 151–159 (2020).

36. El-fatatry, B. M., Ibrahim, O. M., Hussien, F. Z. & Mostafa, T. M. Role of metformin in oxaliplatin-induced peripheral neuropathy in patients with stage III colorectal cancer: randomized, controlled study. Int J Colorectal Dis 33, 1675–1683 (2018).

37. Lin, P. C. et al. N-acetylcysteine has neuroprotective effects against oxaliplatin-based adjuvant chemotherapy in colon cancer patients: preliminary data. Support Care Cancer 14, 484–487 (2006).

38. Milla, P. et al. Administration of reduced glutathione in FOLFOX4 adjuvant treatment for colorectal cancer: effect on oxaliplatin pharmacokinetics, Pt-DNA adduct formation, and neurotoxicity. Anticancer Drugs 20, 396–402 (2009).

39. Lee, S. et al. Potential Mediators of Oxaliplatin-Induced Peripheral Neuropathy From Adjuvant Therapy in Stage III Colon Cancer: Findings From CALGB (Alliance)/SWOG 80702. Journal of Clinical Oncology 41, 1079–1091 (2023).

40. Yehia, R., Saleh, S., El Abhar, H., Saad, A. S. & Schaalan, M. L-Carnosine protects against Oxaliplatin-induced peripheral neuropathy in colorectal cancer patients: A perspective on targeting Nrf-2 and NF-κB pathways. Toxicol Appl Pharmacol 365, 41–50 (2019).

41. Kobayashi, M. et al. Protective effect of the oral administration of cystine and theanine on oxaliplatin-induced peripheral neuropathy: a pilot randomized trial. Int J Clin Oncol 25, 1814–1821 (2020).

42. Motoo, Y., Tomita, Y. & Fujita, H. Prophylactic efficacy of ninjin’yoeito for oxaliplatin-induced cumulative peripheral neuropathy in patients with colorectal cancer receiving postoperative adjuvant chemotherapy: a randomized, open-label, phase 2 trial (HOPE-2). Int J Clin Oncol 25, 1123–1129 (2020).

43. Zhang, R. X. et al. Neuroprotective effect of neurotropin on chronic oxaliplatin-induced neurotoxicity in stage II and stage III colorectal cancer patients: Results from a prospective, randomised, single-centre, pilot clinical trial. Int J Colorectal Dis 27, 1645–1650 (2012).

44. Kuriyama, A. & Endo, K. Goshajinkigan for prevention of chemotherapy-induced peripheral neuropathy: a systematic review and meta-analysis. Supportive Care in Cancer 26, 1051–1059 (2018).

45. Buczyńska, A., Sidorkiewicz, I., Krętowski, A. J. & Adamska, A. Examining the clinical relevance of metformin as an antioxidant intervention. Front Pharmacol 15, 1330797 (2024).

46. Caruso, G., Di Pietro, L., Cardaci, V., Maugeri, S. & Caraci, F. The therapeutic potential of carnosine: Focus on cellular and molecular mechanisms. Current Research in Pharmacology and Drug Discovery 4, 100153 (2023).

47. Grolleau, F. et al. A possible explanation for a neurotoxic effect of the anticancer agent oxaliplatin on neuronal voltage-gated sodium channels. J Neurophysiol 85, 2293–2297 (2001).

48. Hochster, H. S., Grothey, A. & Childs, B. H. Use of Calcium and Magnesium Salts to Reduce Oxaliplatin-Related Neurotoxicity. Journal of Clinical Oncology 25, 4028–4029 (2007).

49. Argyriou, A. A. et al. Clinical pattern and associations of oxaliplatin acute neurotoxicity: a prospective study in 170 patients with colorectal cancer. Cancer 119, 438–444 (2013).

50. Luque-Ceballos, J. C., Rodríguez-Zamora, P., López-Olivos, J. C. & Garzón, I. L. Revisiting the scavenging activity of glutathione: Free radicals diversity and reaction mechanisms. Comput Theor Chem 1227, 114227 (2023).

51. Solana-Manrique, C., Sanz, F. J., Martínez-Carrión, G. & Paricio, N. Antioxidant and Neuroprotective Effects of Carnosine: Therapeutic Implications in Neurodegenerative Diseases. Antioxidants *2022, Vol.* 11, *Page* 848 11, 848 (2022).

52. Aldini, G. et al. Understanding the antioxidant and carbonyl sequestering activity of carnosine: direct and indirect mechanisms. Free Radic Res 55, 321–330 (2021).

53. Ueda, K. et al. Polaprezinc (Zinc L-carnosine) is a potent inducer of anti-oxidative stress enzyme, heme oxygenase (HO)-1 - a new mechanism of gastric mucosal protection. J Pharmacol Sci 110, 285–294 (2009).

54. Anderson, E. J. et al. A carnosine analog mitigates metabolic disorders of obesity by reducing carbonyl stress. J Clin Invest 128, 5280–5293 (2018).

55. Dumitrescu, R., Mehedintu, C., Briceag, I., Purcărea, V. L. & Hudita, D. Metformin-Clinical Pharmacology in PCOs. J Med Life 8, 187 (2015).

56. Wheaton, W. W. et al. Metformin inhibits mitochondrial complex I of cancer cells to reduce tumorigenesis. Elife 3, (2014).

57. Demaré, S., Kothari, A., Calcutt, N. A. & Fernyhough, P. Metformin as a potential therapeutic for neurological disease: mobilizing AMPK to repair the nervous system. Expert Rev Neurother 21, 45–63 (2021).

58. Miao, H. et al. Protective Effects of Vitamin E on Chemotherapy-Induced Peripheral Neuropathy: A Meta-Analysis of Randomized Controlled Trials. Ann Nutr Metab 77, 127–137 (2021).

59. Fu, X. et al. Efficacy of drug interventions for chemotherapy-induced chronic peripheral neurotoxicity: A network meta-analysis. Front Neurol 8, 244755 (2017).

60. Molassiotis, A. et al. Are we mis-estimating chemotherapy-induced peripheral neuropathy? Analysis of assessment methodologies from a prospective, multinational, longitudinal cohort study of patients receiving neurotoxic chemotherapy. BMC Cancer 19, 1–19 (2019).

61. Created in BioRender. Openshaw, M. (2025) https://BioRender.com/e68u958.

