## Supplementary figures for "Anti-oxidative stress therapies reduce chemotherapy-induced peripheral neuropathy in colorectal cancer patients treated with oxaliplatin: a systematic review and meta-analysis"

Salama et al, Supplementary data

*Supplementary file Salama_Supp.xlsx contains:*

**Supplementary Table 1:** Extended summary information for the 29 trials reviewed

**Supplementary Table 2:** Extended information about the 20 studies included in the systematic review and meta-analysis

**Supplementary Table 3:** Summary information for 43 studies reviewed in Peng et al, 2022 that tested a pharmacological intervention for prevention of OIPN, plus reasons trials were excluded from our study.

**Supplementary Table 4: Interventions showing a significant reduction in grade 2 OIPN that were evaluated only by a single study.**

Cycle 6 equates to 2-3 months of chemotherapy; cycle 8 equates to 4-6 months.

| Study and Reference | Intervention tested | Timepoint when OIPN was assessed | P-value | Intervention arm (N) | Placebo/Control arm (N) |
| --- | --- | --- | --- | --- | --- |
| Kobayashi et al.^41^ | Cystine and theanine | Cycle 6 | 0.017 | 14 | 14 |
| Liu et el.^34^ | Gauilongtongluofang | Cycle 6 | 0.04 | 60 | 60 |
| Motoo et al.^42^ | Ninjin’yoeito) | Cycle 8 | <0.01 | 20 | 20 |
| Zhang et al.^43^ | Neurotropin | Cycle 8 | 0.001 | 38 | 41 |

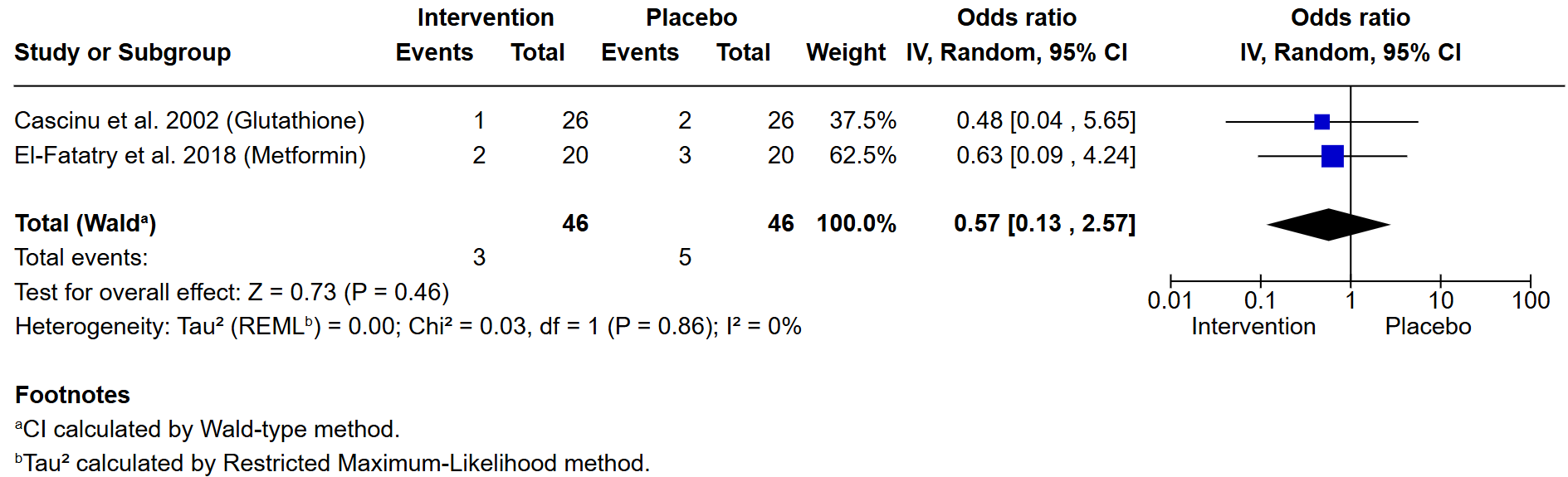

**Supplementary Figure 1. Treatment with anti-oxidative stress drugs glutathione or metformin is not associated with a reduction in CTCAE grade ≥2 peripheral neuropathy after 4-6 cycles of oxaliplatin-based chemotherapy for colorectal cancer.**

Meta-analysis of 2 studies at the mid-point of chemotherapy shows shown no reduction severe of neuropathy *vs.* placebo (P=0.46). The intervention tested in each trial is indicated in parentheses.

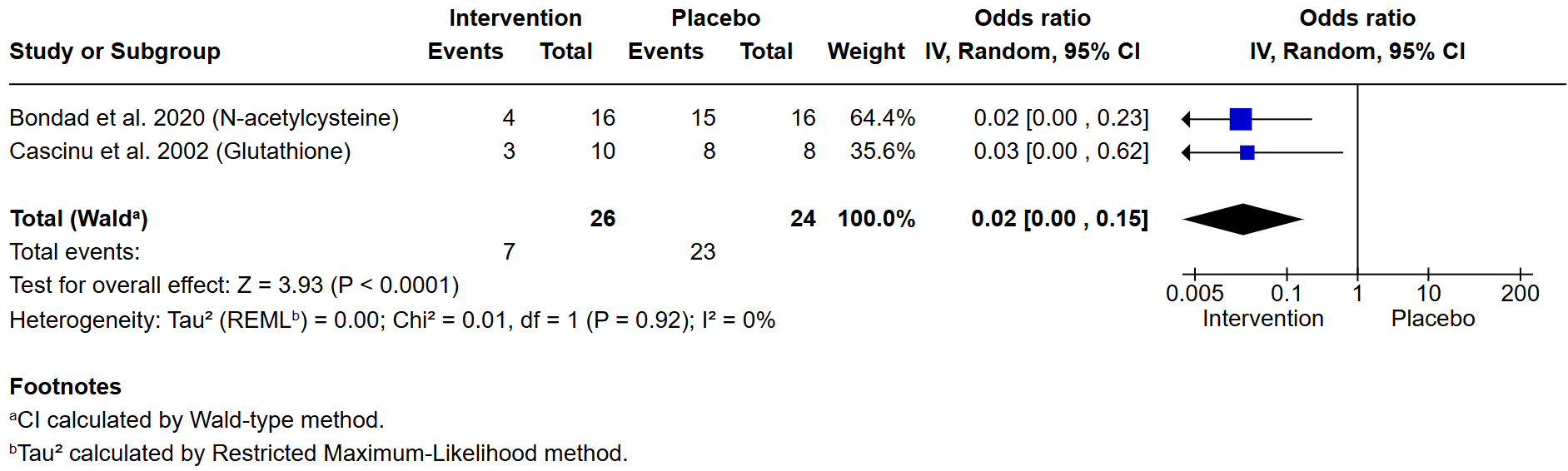

**Supplementary Figure 2. Treatment with anti-oxidative stress drugs n-acetylcysteine or glutathione is associated with a reduction in CTCAE grade ≥2 peripheral neuropathy after 8-12 cycles of oxaliplatin-based chemotherapy for colorectal cancer.** Meta-analysis of 2 studies after 6 months of chemotherapy shows show a highly significant reduction severe of neuropathy *vs.* placebo (P<0.0001). The intervention tested in each trial is indicated in parentheses.
